# The COMBAT-TB Workbench: Making powerful TB bioinformatics accessible

**DOI:** 10.1101/2021.09.23.21263983

**Authors:** Peter van Heusden, Ziphozakhe Mashologu, Thoba Lose, Robin Warren, Alan Christoffels

**Affiliations:** South African Medical Research Council Bioinformatics Unit, South African National Bioinformatics Institute, University of the Western Cape, South Africa; DSI-NRF Centre of Excellence for Biomedical Tuberculosis Research, South African Medical Research Council Centre for Tuberculosis Research, Division of Molecular Biology and Human Genetics, Faculty of Medicine and Health Sciences, Stellenbosch University, Cape Town, South Africa; Africa Centres for Disease Control and Prevention, African Union Headquarters, Addis Ababa W21K19, Ethiopia

## Abstract

Whole Genome Sequencing (WGS) is a powerful method for detecting drug resistance, genetic diversity and transmission dynamics of *Mycobacterium tuberculosis*. Implementation of WGS in public health microbiology laboratories is impeded by a lack of user-friendly, automated and semi-automated pipelines. We present the COMBAT-TB workbench, a modular, easy to install application that provides a web based environment for *Mycobacterium tuberculosis* bioinformatics. The COMBAT-TB Workbench is built using two main software components: the IRIDA Platform for its web-based user interface and data management capabilities and the Galaxy bioinformatics workflow platform for workflow execution. These components are combined into a single easy to install application using Docker container technology. We implemented two workflows, for *M. tuberculosis* sample analysis and phylogeny, in Galaxy. Building our workflows involved updating some Galaxy tools (Trimmomatic, snippy and snp-sites) and writing new Galaxy tools (snp-dists, TB-Profiler, tb_variant_filter and TB Variant Report). The irida-wf-ga2xml tool was updated to be able to work with recent versions of Galaxy and was further developed into IRIDA plugins for both workflows. In the case of the *M. tuberculosis* sample analysis an interface was added to update the metadata stored for each sequence sample with results gleaned from the Galaxy workflow output. Data can be loaded into the COMBAT-TB Workbench via the web interface or via the command line IRIDA uploader tool. The COMBAT-TB Workbench application deploys IRIDA, the COMBAT-TB IRIDA plugins, the MariaDB database and Galaxy using Docker containers (https://github.com/COMBAT-TB/irida-galaxy-deploy).

**Importance:** While the reduction in cost of WGS is making sequencing more affordable in Lower and Middle Income Countries, public health laboratories in these countries seldom have access to bioinformaticians and system support engineers adept at using the Linux command line and complex bioinformatics software. The COMBAT-TB Workbench provides an open source, modular, easy to deploy and use environment for managing and analysing *M. tuberculosis* WGS data and thereby makes WGS usable in practice in the LMIC context.

## Introduction

Tuberculosis (TB) until recently the world’s deadliest infectious disease, infecting an estimated 10 million people in 2019 and killing 1.4 million people(1). Whole Genome Sequencing (WGS) of *Mycobacterium tuberculosis*, the bacterium that causes TB, is increasingly being used, at least in high income countries(2), for species and lineage identification, drug resistance profiling and outbreak investigation. Increasing the use of Whole Genome Sequencing in low and middle income countries (LMICs) requires reducing the cost of both sequencing and bioinformatics analysis of sequencing results and reducing the time and effort involved in going from sequence to analysis results(3).

This cost takes the form of equipment, consumables and expertise. The command line tools whose use is prevalent in bioinformatics (4) require skills not readily accessible outside of specialist labs. On the other hand, web based tools such as TB-Profiler (5) and the NIAID TB Portals(6) are restricted to the analyses provided by their authors and often lack features for bulk analysis. Finally platforms like Galaxy(7), while customisable do not provide a data management and analysis user interface specific for common *M. tuberculosis* analysis tasks.

To address these deficiencies, the computational bacterial analytical toolkit for Tuberculosis research (COMBAT-TB) was developed. The COMBAT-TB Workbench (downloadable from https://github.com/COMBAT-TB/irida-galaxy-deploy) represents a modular and accessible open source workbench for storing and analysing *M. tuberculosis* and other microbial WGS data.

## Results

### Design and implementation

The COMBAT-TB Workbench is built using two main software components: the Integrated Rapid Infectious Disease Analysis (IRIDA) platform (8) for its web-based user interface and data management capabilities and the Galaxy bioinformatics workflow platform for workflow execution. These components are combined into a single easy to install application using Docker container technology.

#### Data management & User interface

The IRIDA platform, a project of the Public Health Agency of Canada (PHAC-NML), is a web application written in Java, that provides a user-friendly web-interface for sequencing data and metadata storage, workflow execution and results visualisation. We adopted IRIDA as the basis of the COMBAT-TB workbench because of its integration with the Galaxy platform, its proven track record in public health bioinformatics (PHAC-NML has analysed over 200,000 pathogen samples using IRIDA (44)) and the fact that it is open source. IRIDA stores sequence samples on disk and sample metadata in a MariaDB database (9). Sequence data is shared between IRIDA and the Galaxy analysis platform (reducing data duplication).

#### Scientific workflows and IRIDA plugins

The Workbench uses Galaxy for its bioinformatics workflow composition and execution. Two workflows, for *M. tuberculosis* sample analysis and phylogeny, were implemented in Galaxy. Building workflows in Galaxy involves connecting Galaxy tools to construct an analysis workflow where the Galaxy tools (also known as tool wrappers) themselves connect command line bioinformatics tools to the Galaxy framework. Building our workflows involved updating some Galaxy tools (Trimmomatic (10), snippy (11) and snp-sites (12)) and writing new Galaxy tools (snp-dists (13), TB-Profiler (5), tb_variant_filter (14) and TB Variant Report (15)). In addition to the work on Galaxy tools, the command line tb_variant_filter and TB Variant Report tools were created as part of the COMBAT-TB project.

The irida-wf-ga2xml tool (16) was updated to be able to work with recent versions of Galaxy and it was used to build IRIDA plugin skeletons. These plugin skeletons were further developed into IRIDA plugins for both workflows, in the case of the *M. tuberculosis* sample analysis involving the addition of an interface between the Galaxy workflow output and the metadata stored for each sequence sample. The *M. tuberculosis* Sample Report and *M. tuberculosis* Phylogeny plugins are hosted in Github repositories (https://github.com/COMBAT-TB/irida-plugin-tb-sample-report and https://github.com/COMBAT-TB/irida-plugin-tb-phylogeny respectively) and deployed into IRIDA as part of COMBAT-TB Workbench deployment process.

#### Deployment

The COMBAT-TB Workbench application (available at: https://github.com/COMBAT-TB/irida-galaxy-deploy) deploys IRIDA, the COMBAT-TB IRIDA plugins, the MariaDB database IRIDA uses for metadata storage and Galaxy using Docker containers (17). The Docker containers are orchestrated using docker-compose (18). This allows the entire Workbench to be installed by users without advanced Linux systems administration knowledge and hides the complexity of the underlying software from the user.

#### Integration with external data storage

Data can be loaded into the COMBAT-TB Workbench via the web interface or via the command line IRIDA uploader tool. This allows data from external storage (for example the storage of a sequencing facility) to be loaded into the Workbench in bulk.

### Comparison to other similar open source software

At the time of writing we found two systems, Innuendo (19) and the IRIDA project (8) that were comparable to the COMBAT-TB Workbench.

Innuendo is a web interface to sequence storage and Nextflow (20) workflow execution aimed at analysis of food-borne pathogens. It is oriented around common tasks in the food-borne pathogens surveillance terrain such as molecular typing of pathogens. The platform is strongly tied to the workflows of a food-borne pathogen surveillance lab, and adding additional species to the analysis system requires modifying the underlying database and adding a wgMLST scheme. As such, Innuendo is addressing a different challenge to the one the COMBAT-TB project is tackling.

IRIDA is the official bioinformatics platform for public health genomics within the Public Health Agency of Canada. It has been in use since 2016 by Canada’s provincial and national public health laboratories for genomic investigations of foodborne disease outbreaks, as part of PulseNet Canada’s foodborne disease surveillance activities. While it is more flexible than Innuendo, as it allows deployment of a wide variety of Galaxy workflows and is not species specific, it is complex to deploy, with the installation guide assuming knowledge of deployment of both Galaxy and the Tomcat Java Servlet system (21). The COMBAT-TB Workbench, by contrast, is straightforward to deploy with a single Linux command.

### Use Case

For the purpose of these analyses we installed the COMBAT-TB Workbench on a virtual machine with 8 virtual CPUs, 32 GB RAM and 3000 GB hard disk space, running Ubuntu 18.04 with Docker version 19.03.14 and docker-compose version 1.27.4.

The COMBAT-TB Workbench user interface is organised around Projects and Analyses. Projects store sequence samples, are associated with a reference genome and allow controlling sharing and access to samples. Samples can be selected from a project and added to a Cart. Once samples are in the Cart, selecting the Cart displays a Workflow selection screen from which Analyses can be started (Figure 2):

**Figure 1.**
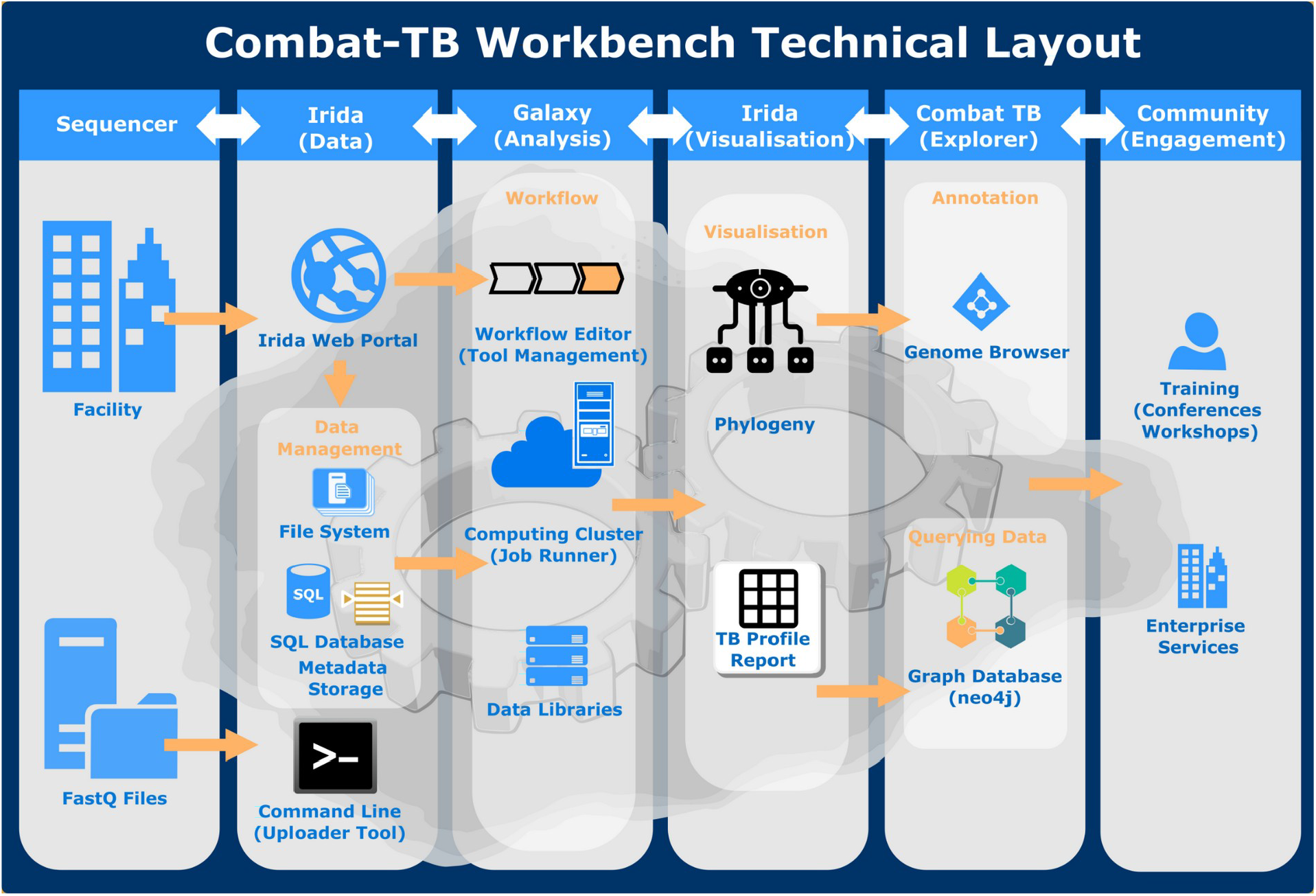
COMBAT-TB Workbench Technical Layout

**Figure 2.**
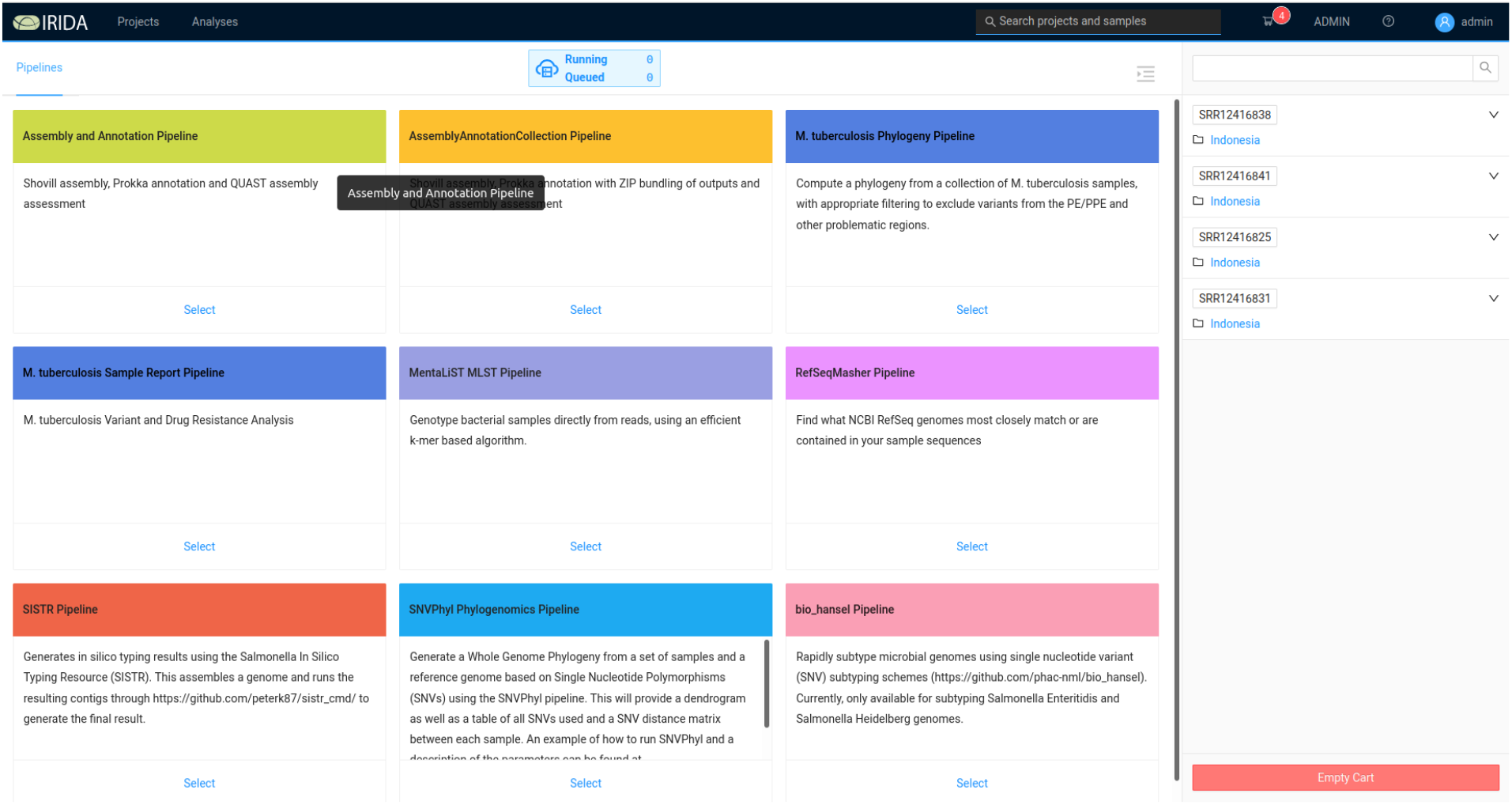
COMBAT-TB Workbench: Workflow selection screen

Selecting one of the Workflows allows the workflow parameters to be set and the analysis to be started. Workflows can either be per-sample, in which case a workflow instance is started for each sample, or multi-sample (for example phylogenies) in which case the samples are analysed as a group. Once a workflow has been executed it creates a new entry visible via the Analyses interface. This interface allows monitoring the status of workflow execution and visualization of workflow analysis results.

#### Analysis of data from MDR *M. tuberculosis* in Indonesia

Tania et al (22) collected *M. tuberculosis* samples from 30 patients with confirmed pulmonary tuberculosis treated in four hospitals in the Western region of the Indonesian island of Java. Phenotypic drug susceptibility testing (DST) was performed on the cultured samples and DNA was isolated, sequenced and deposited in the NCBI Sequence Read Archive (SRA) with BioProject access number PRJNA633244. Thirty samples were retrieved from EBI European Nucleotide Archive (ENA) and uploaded to the COMBAT-TB Workbench using the irida-uploader (23) command line tool. Per sample quality control was performed automatically (using FastQC(24)) after sample upload. Uploading and quality control took 9 and 3 minutes respectively.

The inferred ancestral *M. tuberculosis* reference genome produced by Comas et al (25) was downloaded from Zenodo (26) and uploaded to the COMBAT-TB Workbench as the default reference genome for the sequencing project. While the H37Rv reference genome can be used, we used the *M. tuberculosis* inferred ancestral reference as Goig et al (27) have previously shown that this genome is equidistant, in terms of sequence variants, from all known *M. tuberculosis* lineages and thus provides a superior reference to H37Rv (NC_000962.3) for variant calling, especially if that variant calling is going to be used for phylogeny construction.

#### Per sample reporting

The *M. tuberculosis* Sample Report pipeline (see Materials and Methods) was run on all 30 samples. Detailed output from this pipeline is made available in a per-sample analysis report (Supplementary Figure 1) that includes a full report on variants identified in the sample (annotated using information from the COMBAT-TB NeoDB (28)), drug resistance prediction (from TB-Profiler) and quality control information on read mapping.

The pipeline updated the metadata associated with each sample, with the result that after pipeline completion each sample was annotated with percentage of reads mapped to genome, drug resistance prediction, drug resistance associated variants and lineage assigned (Figure 3).

**Figure 3:**
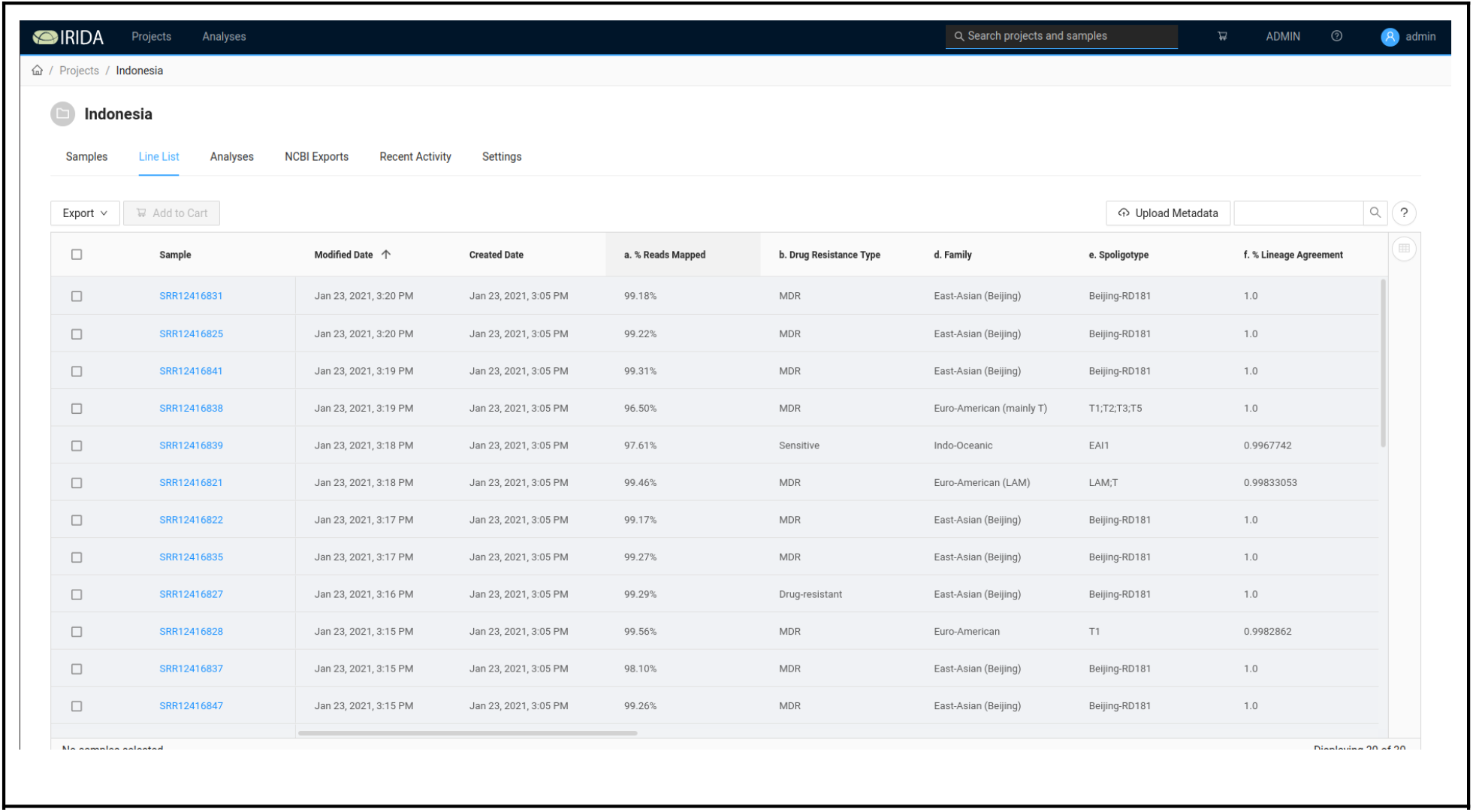
Sample Line List

Examining the read mapping outputs shows that only 9.63% and 1.47% of reads from samples SRR12416824 and SRR12416842 respectively mapped against the *M. tuberculosis* genome. Further investigation with kraken2 (29) using the Standard database from 14 April 2020 showed that the majority (63.55%) of reads from SRR12416824 were classified as belonging to the *Mycobacterium avium* complex (MAC) and the majority of reads (75.39%) from SRR12416842 were classified as the non-tubercular mycobacterium *Mycolicibacterium fortuitum*. These samples were thus excluded and not used in subsequent analyses.

As shown in Table 1, the results were broadly concordant with those found by Tania et al. Some small differences are likely a result of different versions of the TB-Profiler software used. Without our own analysis, updating the TB-Profiler version from 2.8.4 to version 3.0.6 reduced the discordance between streptomycin resistance predicted by the COMBAT-TB Workbench and that predicted by MGIT by 2 samples. This illustrates the evolution of drug resistance prediction from WGS data. As noted previously, the COMBAT-TB Workbench pipeline uses snippy for mapping and variant calling and applies TB-Profiler only to the results of the mapping step. Our results are, however, wholly concordant with running the complete TB-Profiler pipeline.

**Table 1:** Drug Resistance predictions from COMBAT TB Workbench & Tania et al

| Drug | Concordant<br>MGIT | Discordant<br>MGIT<br>Resistant | Discordant<br>MGIT<br>Susceptible | Concordant<br>Tania et al<br>TB-Profiler | Discordant<br>Tania et al<br>Resistant |
| --- | --- | --- | --- | --- | --- |
| Isoniazid | 24/28<br>(85.71%) | 2 | 2 | 23/28<br>(82.14%) | 1 |
| Rifampicin | 27/28<br>(96.43%) | 0 | 1 | 27/28<br>(96.43%) | 0 |
| Ethambutol | 17/28<br>(60.71%) | 0 | 11 | 18/28<br>(64.29%) | 1 |
| Streptomycin | 25/28<br>(85.71%) | 2 | 1 | 23/28<br>(82.14%) | 2 |

The lower degree of concordance found between WGS and MGIT resistance testing for ethambutol can be explained by the presence of variants impacting amino acid 306 of the embB gene in 7 out of the 11 samples that were reported as resistant by WGS but susceptible by MGIT. These variants (M306V in 2 and M306I in 5) lead to a change in the MIC of ethambutol (30).

#### Phylogeny on all samples

The 28 samples that previously passed quality control were selected in the web interface and submitted to the *M. tuberculosis* phylogeny pipeline (https://github.com/COMBAT-TB/irida-plugin-tb-phylogeny). This pipeline computes a Maximum Likelihood Phylogeny using the single nucleotide variants identified in each sample (Figure 4).

**Figure 4:**
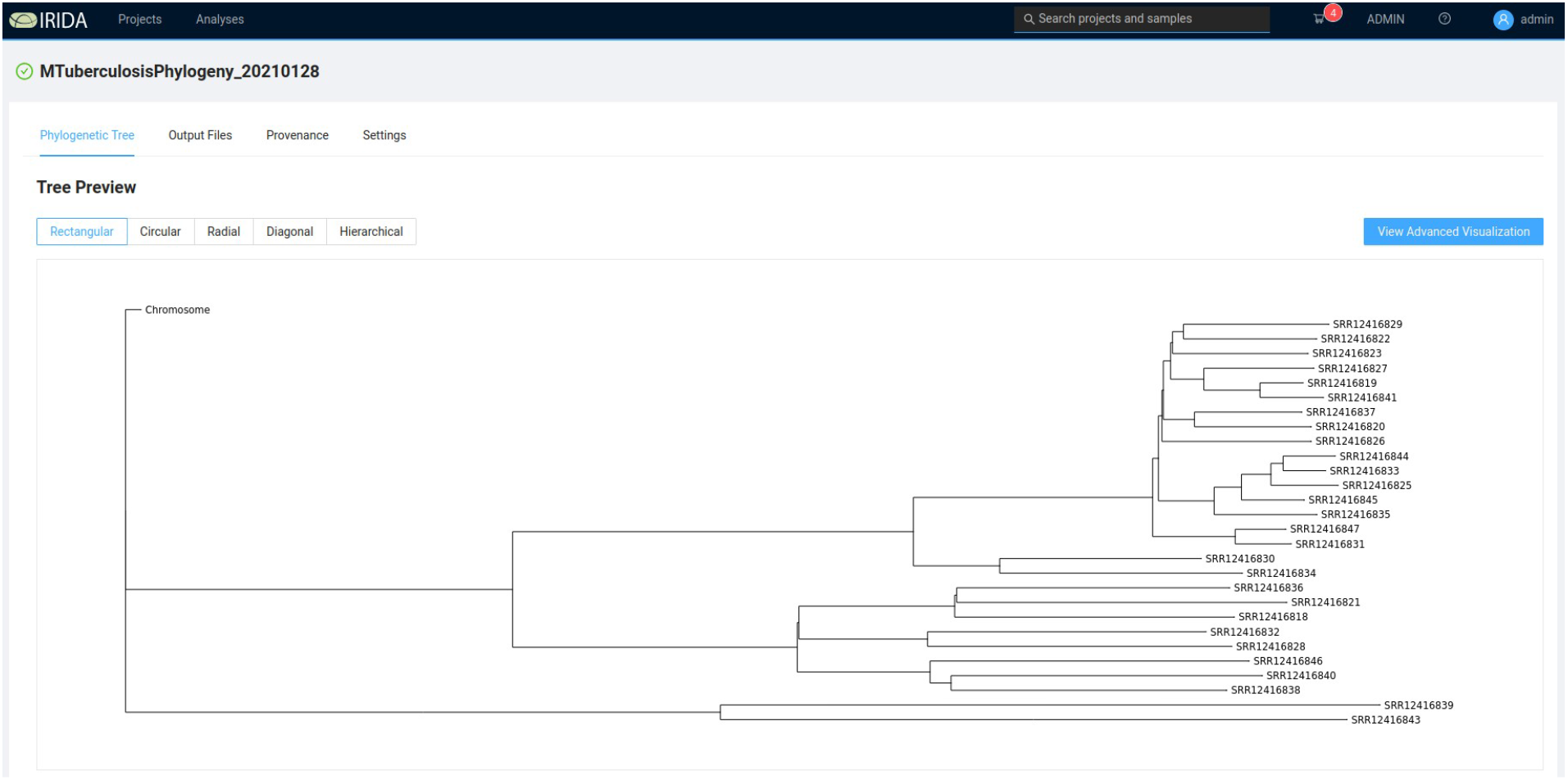
COMBAT TB Workbench phylogeny viewer

The IRIDA Advanced Visualization view (Figure 5) allows metadata from the sequence analysis project’s sequencing store to be associated with tips (i.e. samples) in the phylogeny view. In addition to the metadata computed by the *M. tuberculosis* Sample Report pipeline, an Excel spreadsheet was generated associating each sample ID with the sample IDs and hospital collection sites identified in the Tania et al. paper. This spreadsheet was used to load additional metadata into the IRIDA project.

**Figure 5:**
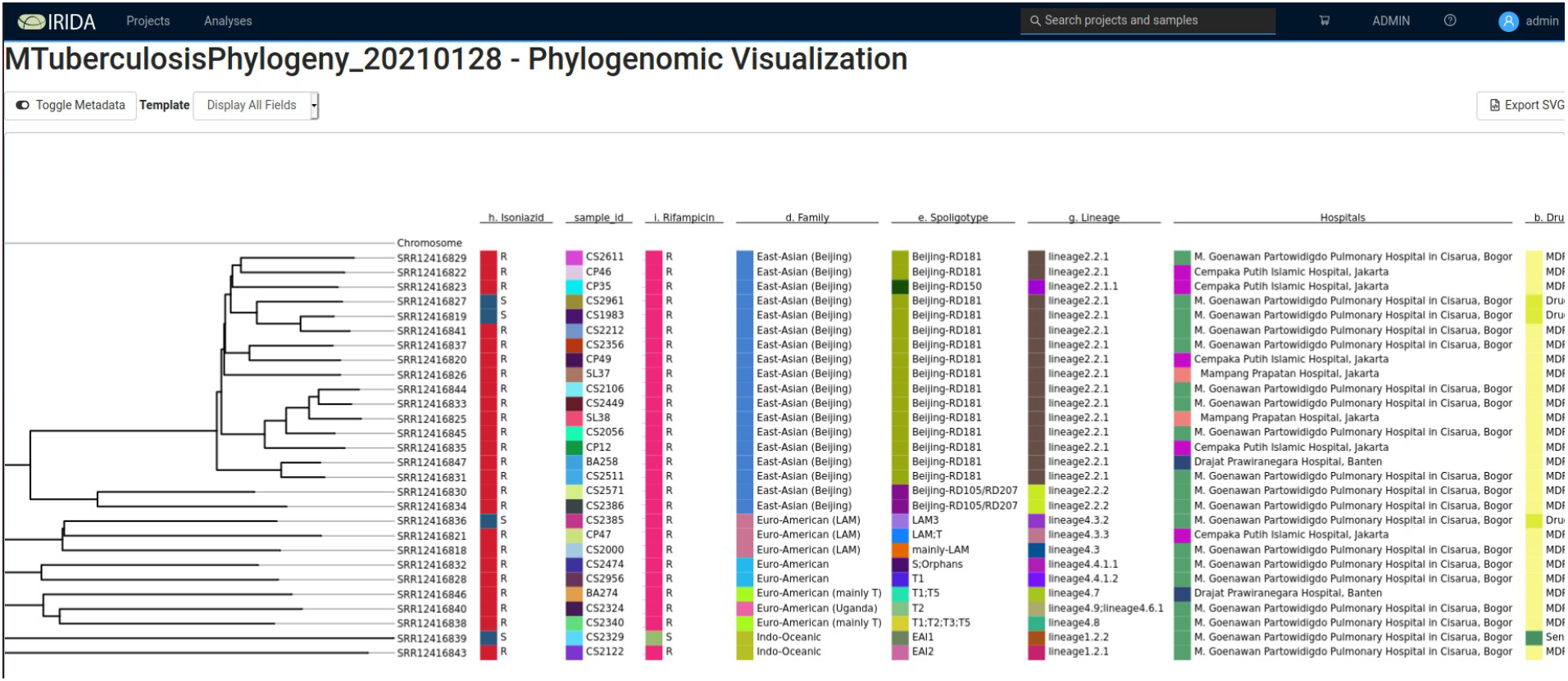
Advanced Phylogeny Visualisation with Metadata columns

The distance between hospital sites varied from 10km (Mampang Prapatan Hospital, Jakarta to Cempaka Putih Islamic Hospital, Jakarta) to 107km (Drajat Prawiranegara Hospital, Serang to M. Goenawan Partowidigdo Pulmonary Hospital, Bogor). When visualised this way it was apparently that there was no clear relationship between phylogenetic relationship (and thus lineage) and hospital site, illustrating that the outbreaks occurring in the region were circulating in areas broader than hospital catchment areas.

#### Analysis of runtimes

The runtimes of the steps in the analyses are listed in Table 2. Galaxy scheduled the execution of analysis steps in parallel when data dependencies allowed. The similarities of the runtimes between the sample processing and the phylogeny pipelines are to some extent due to the fact that IRIDA currently starts all analyses from sequence reads rather than from assembled genomes. This limitation is currently being addressed by the IRIDA development community. (31)

**Table 2:**
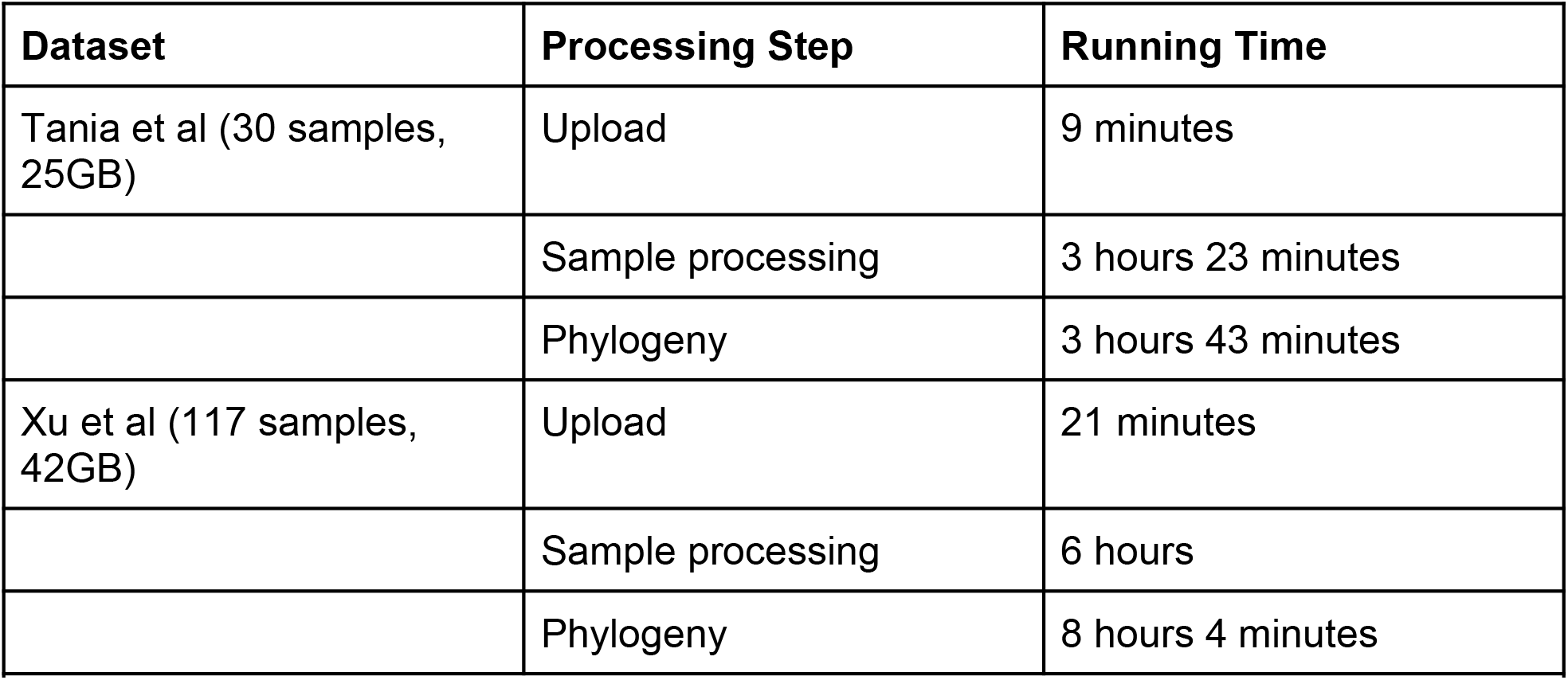
Runtime of analyses in COMBAT TB Workbench

| Dataset | Processing Step | Running Time |
| --- | --- | --- |
| Tania et al (30 samples, 25GB) | Upload | 9 minutes |
|  | Sample processing | 3 hours 23 minutes |
|  | Phylogeny | 3 hours 43 minutes |
| Xu et al (117 samples, 42GB) | Upload | 21 minutes |
|  | Sample processing | 6 hours |
|  | Phylogeny | 8 hours 4 minutes |
| Table 2: Runtime of analyses in COMBAT TB Workbench |  |  |

#### Analysis of data from *M. tuberculosis* in Spain

Xu et al (32) collected *M. tuberculosis* samples from 117 patients and analysed them to understand dynamics of TB transmission in the Valencia Region, in Spain. While the key element of their analysis is identifying individual transmission patterns, their work also provides a convenient larger dataset to examine the performance of the COMBAT-TB Workbench

Uploading of the 117 samples took 21 minutes. Per sample analysis of the 117 samples took a total of 6 hours. A phylogeny was computed using the samples. Phylogeny generation took just over 8 hours and visualisation with the advanced phylogeny viewer revealed clustering by *M. tuberculosis* lineage (Figure 7) as expected. Unfortunately Xu et al did not include the output of their phylogeny construction in their results, so a direct comparison of the computed phylogenetic trees is not possible.

**Figure 7:**
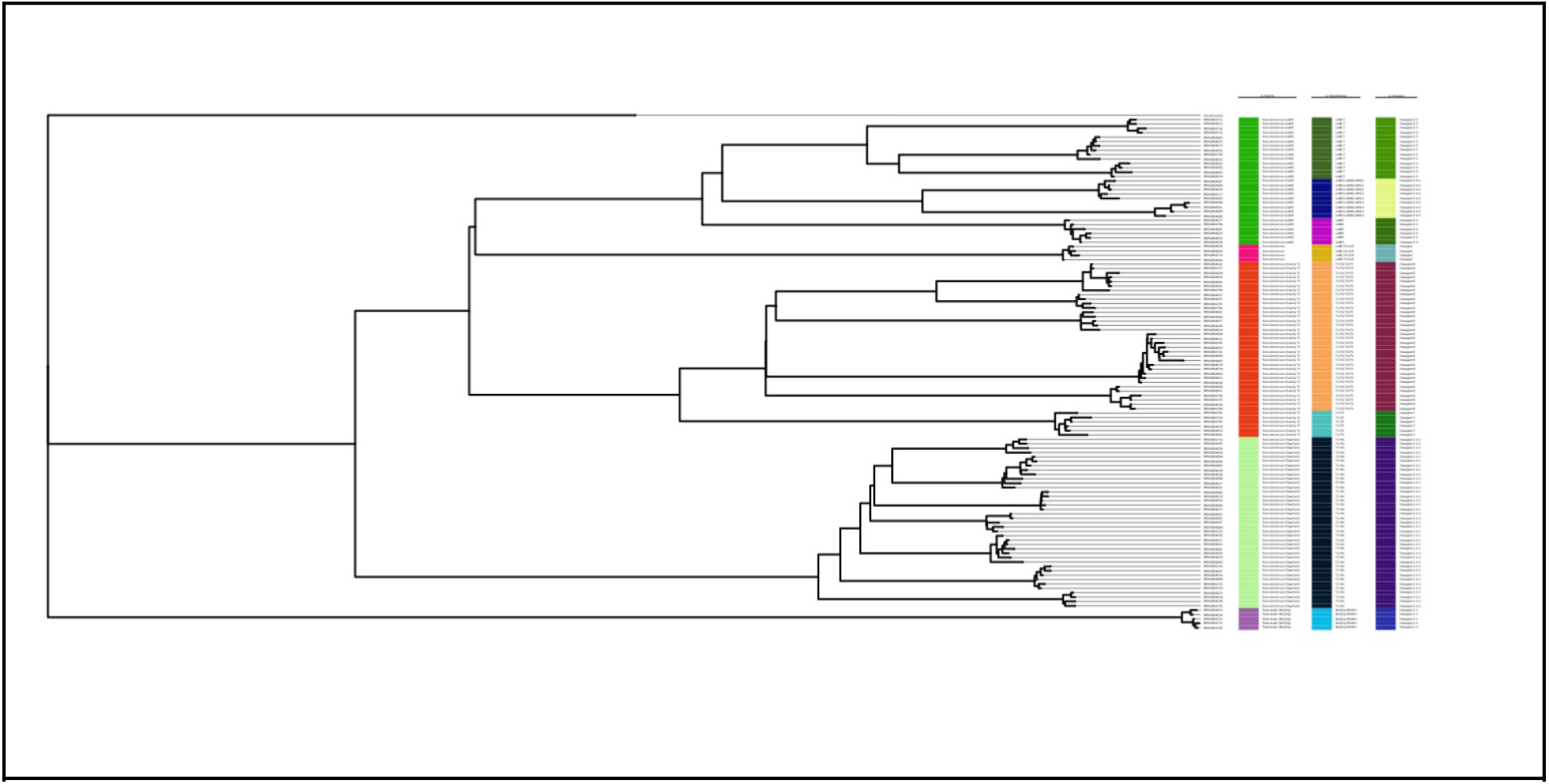
Advanced Phylogeny viewer with Xu et al data (metadata shows lineage)

## Discussion

The COMBAT-TB Workbench makes routine *M. tuberculosis* WGS sample storage and bioinformatics analysis accessible in an extensible framework. The IRIDA platform on which it is built allows pipelines to be added as needed, and the use of Docker container technology means that installation on a machine (with the required Docker and docker-compose software) is a straightforward process not requiring advanced systems administration skills.

By making this project, developed using open source best practices, available the cost of use of WGS can be reduced, increasing opportunities for *M. tuberculosis* WGS deployment in low and middle income countries. As the capabilities of the underlying IRIDA and Galaxy platforms continue to evolve, the COMBAT-TB workbench will naturally acquire additional features in addition to the features being added by the authors.

## Materials and Methods

### Implementation of the COMBAT-TB Workbench

IRIDA and Galaxy are both server environments, and IRIDA relies on a MariaDB database. The COMBAT-TB Workbench executes each of these servers in Docker containers and the execution of the COMBAT-TB Workbench as a whole is orchestrated using docker-compose. The COMBAT-TB Workbench is deployed on any machine that runs Docker and docker-compose by fetching its code from Github (https://github.com/COMBAT-TB/irida-galaxy-deploy) and running a single command (“docker-compose up --build -d”). Updates to the Workbench are similarly applied using a single command. The COMBAT-TB Workbench updates its IRIDA plugins from their repositories on Github on startup.

#### Implementation of the *M. tuberculosis* Sample Report pipeline

The TB sample report workflow starts by running Trimmomatic (v. 0.38.1) with the Sliding Window trimmer, truncating reads when the average quality within a four base window drops below quality score 30, followed by the Minimum length trimmer that discards reads shorter than 20 bases. Only reads which remain in read pairs after quality trimming are used in subsequent analysis.

After quality trimming, sequence reads are mapped to a user supplied reference genome. The workflow requires a genome with the same coordinate scheme as the *M. tuberculosis* H37Rv reference (RefSeq NC000962). Mapping and variant calling is done using snippy (v. 4.4.5), a Perl-based pipeline that combines the bwa-mem (v. 0.7.17) (33) mapper and the freebayes (v. 1.3.2) (34) variant caller. Snippy has been shown (35) to produce good quality variant calls in *M. tuberculosis* when run with default parameters. Samtools (v. 1.9) (36) flagstat is run to provide an overview of statistics from the mapping process.

After variant calling, variants are annotated with SnpEff (v. 4.3) (37) using the H37Rv reference genome annotation. Variants are then filtered using tb_variant_filter (v. 0.1.3). While snippy performs quality based filtering of the variants it predicts, this tool offers a variety of filtering options commonly used in *M. tuberculosis* variant filtering. In our workflow it filters out variants in the PE/PPE gene regions (38), in the repetitive and insertion section regions identified by UVP (39) and those with lower than 30 supporting reads or within 5 base pairs of an indel. These filtering options like all tool options in the workflow can optionally be altered by the user.

In parallel to the SnpEff annotation and variant filtering steps, the mapped reads are provided to TBProfiler (v. 2.8.4) which performs its own variant calling and lineage and drug resistance prediction.

Finally the filtered, annotated variants and the TBProfiler results are fed to tb_vcf_report (v. 0.1.7) which produces a report further annotated with information from the COMBAT-TB eXplorer database (28) in both text and HTML formats.

The final reports provided to the user are the variant reports from tb_vcf_report, text and JSON format reports from TBProfiler, variants in VCF format from SnpEff and mapping statistics from samtools flagstats. These reports include both user readable and raw data suitable for further downstream analysis. The metadata stored in the sample line list is updated with mapping %, *M. tuberculosis* lineage and spoligotype information and drug resistance information.

#### Implementation of the *M. tuberculosis* Phylogeny pipeline

The workflow in the phylogeny module starts with quality filtering of samples using fastp (v. 0.19.5) (40) with default settings. The filtered reads are then aligned to the user provided reference using snippy and predicted variants are filtered with tb_variant_filter as described above. In addition, only single nucleotide variants (SNVs) are retained as the phylogeny software used in the workflow cannot extract meaningful information from indels.

For each sample, the identified sequence variants are inserted into the reference genome, yielding one sequence per sample, of the same length as the reference but with variants from the sample inserted. These are concatenated into a multiple sequence alignment (MSA), which is used as input to snp_dists (v. 0.6.3). Variant and constant sites are identified using snp_sites (v. 2.5.1) and the multiple sequence alignment is filtered to retain only variant sites. A phylogeny is then built using IQ-TREE (v. 1.5.5.1) (41–43).

The final report includes the SNP distance matrix and (Newick format) tree. The tree is also visualised in a phylogeny viewer and can also be displayed with associated sample metadata.

The SNP distance matrix, the phylogeny and the filtered variants (in VCF format) are presented to the user as output.

## Data Availability

The COMBAT-TB Workbench open source software released under the Apache 2.0 license.

https://github.com/COMBAT-TB/irida-galaxy-deploy

https://github.com/COMBAT-TB/irida-plugin-tb-sample-report

https://github.com/COMBAT-TB/irida-plugin-tb-phylogeny

## Funding

This work was supported by The South African Research Chairs Initiatives of the Department of Science and Technology and National Research Foundation of South Africa grant UID 64751, and the South African Medical Research Council flagship programme MRC-RFA-UFSP-01-2013/ COMBAT-TB.

## Supplemental Figures

**Supplementary Figure 1:**
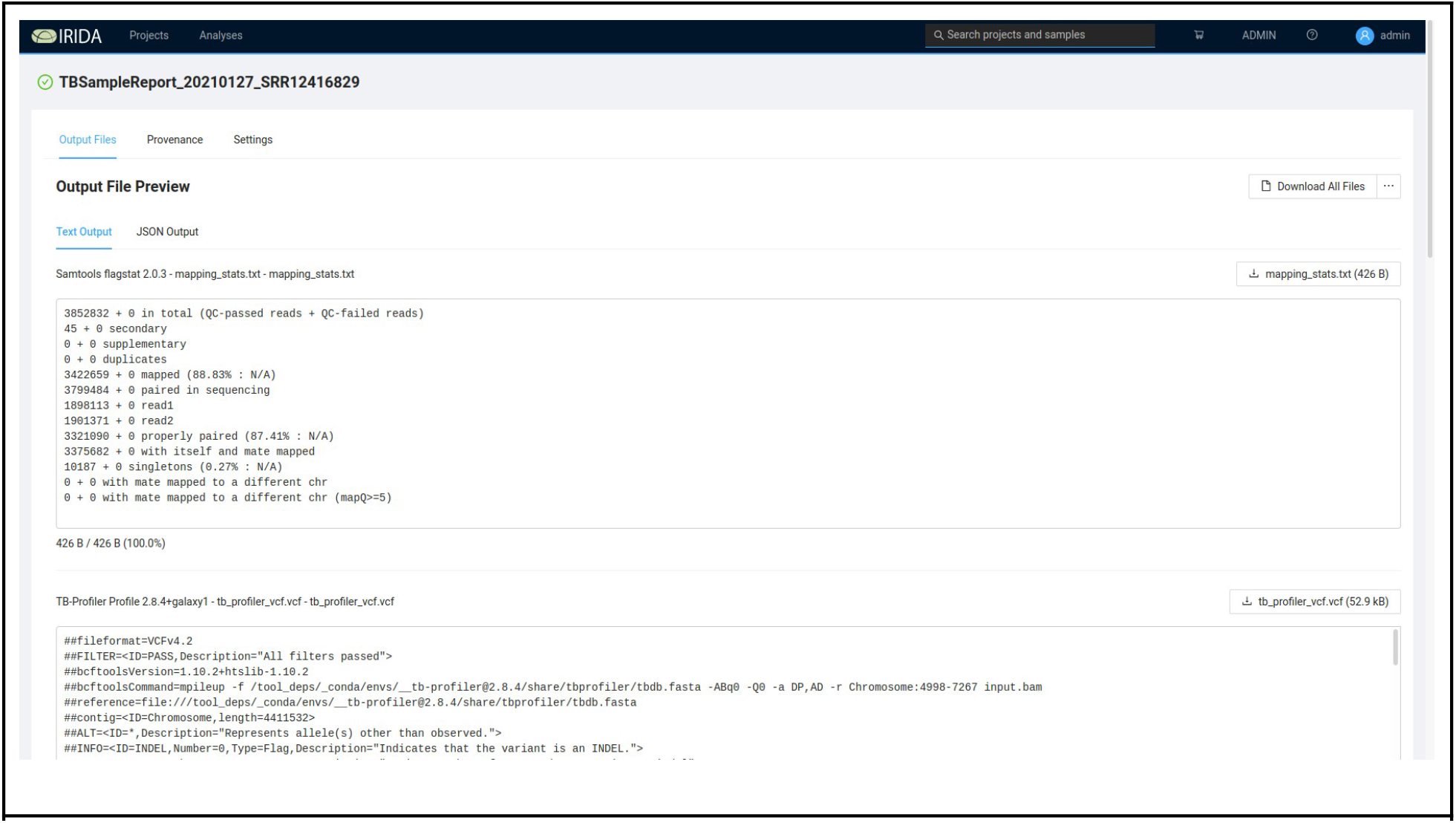
Drug Resistance Report

## Notes

### Competing Interest Statement

The authors have declared no competing interest.

### Summary of Updates

Correct author name, correct corresponding author details.

